# Rethinking cholera diagnostic test performance, interpretation and evaluation: a field-based latent-class analysis in Bangladesh

**DOI:** 10.1101/2024.11.19.24317512

**Authors:** Javier Perez-Saez, Taufiqur Rahman Bhuiyan, Sonia T Hegde, Ishtiakul Islam Khan, Md Taufiqul Islam, Zahid Hasan Khan, Mohammad Ashraful Amin, Juan Dent Hulse, Shakeel Ahmed, Mamunur Rashid, Rumana Rashid, Md Zakir Hossain, Ashraful Islam Khan, Firdausi Qadri, Andrew S Azman

**Author notes:** co-first authorship (Javier Perez-Saez, Taufiqur Rahman Bhuiyan, Sonia T Hegde). co-senior authorship (Firdausi Qadri, Andrew S Azman).

## Abstract

**Background:** Accurate and reliable diagnostics, including rapid diagnostic tests (RDTs), are critical components of cholera control programs, though their performance has varied greatly across studies. While poorly understood, this variability may be due to the reference assay choice, patient-level and/or sampling characteristics, which hinder test result interpretation and performance evaluation.

**Methods:** We enrolled all suspected cholera cases seeking care at two healthcare facilities in Sitakunda, Bangladesh over 19 months. All stool samples were tested with the Cholkit RDT, and a subset by PCR and culture. Test performance was estimated using a latent-class Bayesian framework accounting for imperfect test performance, incomplete PCR and culture testing, and time-varying changes in cholera incidence. Patient-level (including age, antibiotic use) and sampling (season, testing delays) factor effects were estimated, and simulations were used to assess the bias in RDT performance estimates when using traditional reference assays.

**Findings:** We enrolled 3,744 suspected cases, 692 of whom were RDT-positive. Among the RDT-positives, 573 were PCR-positive and 450 culture-positive. For RDT, PCR and culture, we estimated a sensitivity of 93.5% (95% Credible Intervals, CrI: 91.3-95.4), 90.3% (88.4-92.1), and 73.7% (70.8-76.5), and a specificity of 97.3% (96.7-97.8), 97.2% (96.6-97.8), and 100% (culture specificity assumed perfect), respectively. We found that younger age (≤ 5), antibiotic use, and testing delays decreased culture sensitivity, but RDT performance remained relatively constant. The RDT positive predictive value ranged from <15% in children <5 years to >80% in adults, varying greatly across seasons. Simulations demonstrated underestimation of RDT sensitivity and specificity in low and high cholera prevalence settings, respectively, when evaluated against PCR or culture.

**Interpretation:** Our results shed light on the potential mechanisms leading to heterogeneous cholera RDT performance estimates in previous studies, including the use of culture as a reference assay. Across various patient and sampling characteristics, Cholkit RDT had high performance in this cholera-endemic setting, supporting its use for cholera surveillance and control. Accounting for epidemiologic context is crucial both for individual-level clinical test interpretation, and for the future evaluation of diagnostics like RDTs.

**Funding:** The work was supported by the Bill & Melinda Gates Foundation (INV-021879).

## Introduction

Surveillance is a key pillar in the global effort to eliminate cholera, a disease that still kills an estimated 95,000 people annually.^1,2^ National surveillance systems have largely relied on counting acute watery diarrhea (AWD) cases attending health facilities, which is known to have poor specificity (8.1–43.1%; high probability of false positives) due to the number of other pathogenic causes of AWD.^3,4^ Such difficulties in identifying true cholera cases limits our ability to rapidly detect and confirm cholera outbreaks, which in turn can have impacts on population-level disease control efforts and clinical management.^5^

In an effort to improve true cholera case detection, the Global Taskforce for Cholera Control (GTFCC) published recommendations for national cholera surveillance programs and outbreak responses in 2024,^6^ highlighting the role of rapid diagnostic tests (RDTs) in tracking cholera incidence and rapidly identifying cholera outbreaks. To help broaden the production of and trust in cholera RDTs, the Foundation for Innovative New Diagnostics (FIND) developed a target product profile focused on cholera RDTs for surveillance, setting minimum standards for test accuracy.^7,8^ Despite being commercially available for over a decade, RDT uptake by public health programs has been limited in part due to a lack of consistency in product quality ^9^ and unexplained variability in performance results during field studies^10,11^ (meta-analysis: sensitivity range 66%-100%, specificity range 47%-96%^12^). Factors such as sample collection and processing, reference assays and patient-level factors (including age, antibiotic use and severity) likely shape this wide variability in test performance estimates. While these advances in guidance on RDTs and the increased availability will hopefully lead to improved RDT use, a robust understanding of test performance is also needed to support regulatory authorities and public health officials in deciding how to appropriately use these tests in practice.

RDT evaluation relies on two primary laboratory techniques used to confirm cholera: culture and Polymerase Chain Reaction (PCR).^13 14^ Many cholera-affected countries, however, still lack the capacity to use these systematically and these assays have imperfect performance. Culture, which relies on growing the bacteria *V. cholerae* O1/O139, is highly specific but has moderate to low sensitivity due to variability in the bacterial load of cases, sample storage and transport conditions, patient antibiotic use and phage predation.^15–17^ Alternatively, PCR, which relies on gene amplification, tends to have higher sensitivity and high, though imperfect, specificity.^18^ Without a perfect ‘gold standard’ reference assay, estimates of diagnostic field performance can be biased. Even so, as shown previously, the parallel use of multiple imperfect diagnostic tests combined with latent class statistical models can act as a solution and estimate the true performance of each test.^19–21^

Across pathogens, the interpretation of a diagnostic test result is known to not only depend on the accuracy of the test (sensitivity and specificity), but the pretest probability of disease,^22^ which can vary by geography, time, and population subgroups. Appropriate interpretation of cholera diagnostics, notably RDTs, is cardinal to the successful implementation of cholera control measures: for deciding whether to implement interventions, among whom they should be distributed, and the speed at which this should be done. Robust estimates of the positive and negative predictive values of cholera RDTs, and how they may vary across the year and by age group, are needed to inform practitioners how to interpret test results, thereby improving population-level decision making.

Here, we capitalized on unique study data from a cholera-endemic community in Bangladesh, where multiple diagnostic tests were performed in parallel along with detailed clinical surveillance data. We aimed to quantify cholera RDT, culture and PCR performance in this field setting and to assess how patient-level and contextual factors affect performance. To gain new insights into the practical use of cholera diagnostic tests, we developed latent class models to estimate the accuracy of multiple cholera diagnostics, describe how it differs by sample and patient characteristics, and illustrate how the interpretation of test results can vary across seasons and populations. Finally, we demonstrated through simulations how the choice of reference assay can bias estimates of test performance, helping to explain heterogeneity in previous estimates and providing guidance for future diagnostic evaluation study designs.

## Methods

### Study design and setting

We conducted enhanced clinical surveillance in the Sitakunda sub-district of Chattogram, Bangladesh at the two public health facilities that serve as the primary sites for diarrheal disease and cholera surveillance for the Bangladesh Directorate General for Health Services in Sitakunda: the Sitakunda sub-district hospital (Upazila Health Complex) and the Bangladesh Institute for Tropical Infectious Diseases (BITID).23 Sitakunda covers an area of approximately 500 km2 and has a population of 383,000 people.24 From January 24, 2021 to August 31, 2022, we attempted to enroll all suspected cases ≥1 years old presenting with non-bloody, acute watery diarrhea (3 or more loose non-bloody stools in the 24-hours preceding the visit) at the in-patient and out-patient wards of both facilities. We obtained written informed consent, administered a short, structured questionnaire, and collected a stool (or rectal swab) specimen for laboratory analyses from all participants.

### Laboratory testing

We tested each participant’s fecal sample on site with the CholKit Rapid Diagnostic Test (Incepta, Dhaka, Bangladesh) following manufacturer instructions.^25^ Samples were then placed in Cary Blair media and on Whatman 903 filter paper for subsequent lab testing at icddr,b in Dhaka.^26^ All RDT positive samples were tested by conventional stool culture and end-point PCR (for ctxA and rfb genes from filter paper).^18^ Briefly, for culture, stool was directly streaked onto selective TTGA (taurocholate-tellurite gelatin agar) plates, and plates were incubated overnight at 37°C. Colonies morphologically consistent with *V. cholerae* were tested for agglutination reactions with monoclonal antibodies specific to *V. cholerae* serovar O1 (Ogawa or Inaba) and O139. We additionally tested a random subset of nearly half of the RDT negative samples (45.6%) by PCR. All diagnostic test results were used in the latent class model.

### Univariate tests

We performed univariate analyses on various participant and sampling characteristics to assess differences between those who were RDT-positive and RDT-negative. This included Pearson’s chi-squared tests, Fisher’s exact tests, and Wilcoxon rank sum tests depending on the variable type.

### Latent class model

To infer the performance of the Cholkit RDT, PCR, and culture for cholera diagnosis, we developed a Bayesian modeling framework that accounted for the lack of a gold standard assay, partial testing within our sampling protocol (only roughly half of RDT-negative samples were tested with PCR, and none with culture), as well as age-specific changes in the underlying proportion of cholera cases among AWD during the study period. Priors for the sensitivity and specificity of each test were based on published estimates in a joint evaluation of Cholkit with culture and PCR.^25^ We used the Stan programming language to draw samples from the posterior distribution.^27^ We assessed convergence both visually and through the Rhat statistic and evaluated model fit through posterior retrodictive checks of weekly test counts.

Full details are given in the Supplementary Material section S1.

### Covariate effects on test performance

We assessed the effect of participant characteristics (age, antibiotic use prior to hospitalization) and sampling details (season, delay from sample to laboratory testing) on test performance. We incorporated these variables as covariates in our estimates of test sensitivity and specificity, accounting for possible confounding between variables following our assumptions on the causal links between variables (cf. Directed Acyclic Graphs in Supplementary Figure S1). For antibiotic use, we made the distinction in the main analysis between antibiotics recommended by the GTFCC for cholera treatment (those that typically lack resistance and are effective),^28^ and other or no antibiotics. We conducted sensitivity analyses considering any antibiotic use, and all antibiotics that are known to be effective against *V. cholerae* regardless of known resistance patterns. A full list of covariate effects and regression equations controlling for possible confounding are given in Supplementary Table S1. To obtain posterior estimates along covariate levels we post-stratify covariate-specific results using proportions in our sample (for instance the distribution among age classes for the effect of antibiotic use).

### Simulation of RDT evaluation scenarios

We conducted a simulation study to quantify the biases in RDT (or other diagnostic) evaluations that may result from the choice of study setting due to differences in the underlying cholera prevalence and the imperfect nature of reference assays. We simulated test results by taking the mean estimates of test sensitivity and specificity in this study as references and performed 100 simulations of test outcomes for 300 participants. Each set of simulations was done for values of cholera prevalence among AWD cases ranging from 1% to 99%. We then computed the corresponding estimates of RDT sensitivity and specificity taking three commonly used reference assays: PCR, culture and a PCR-culture composite.

Following previous studies, the composite reference assay considered a participant to be positive if either the PCR or culture result was positive, and negative if both PCR and culture results were negative.^21^

Code and data to reproduce analyses from this paper will be available at https://github.com/HopkinsIDD/rethinking_cholera_diagnostics/.

## Results

From January 24, 2021 to August 31, 2022, 3,744 participants were enrolled into clinical surveillance at the two healthcare facilities mandated to treat cholera in the sub-district of Sitakunda (Table 1). Among the suspected cases, 18.5% (692/3,744) of participants tested positive by the Cholkit RDT. Among the RDT-positive samples, 82.8% also tested positive by PCR, and 65.0% by culture. Among RDT-negative samples that were further tested by PCR, 8.0% were PCR-positive (111/1,391). Positivity rates for all three tests varied significantly between age groups, with children below 5 having the smallest proportion of PCR-positives among those testing positive by RDT (53.8%, 43/80) (Figure 1, Supplementary Figure S2).

**Table 1.**
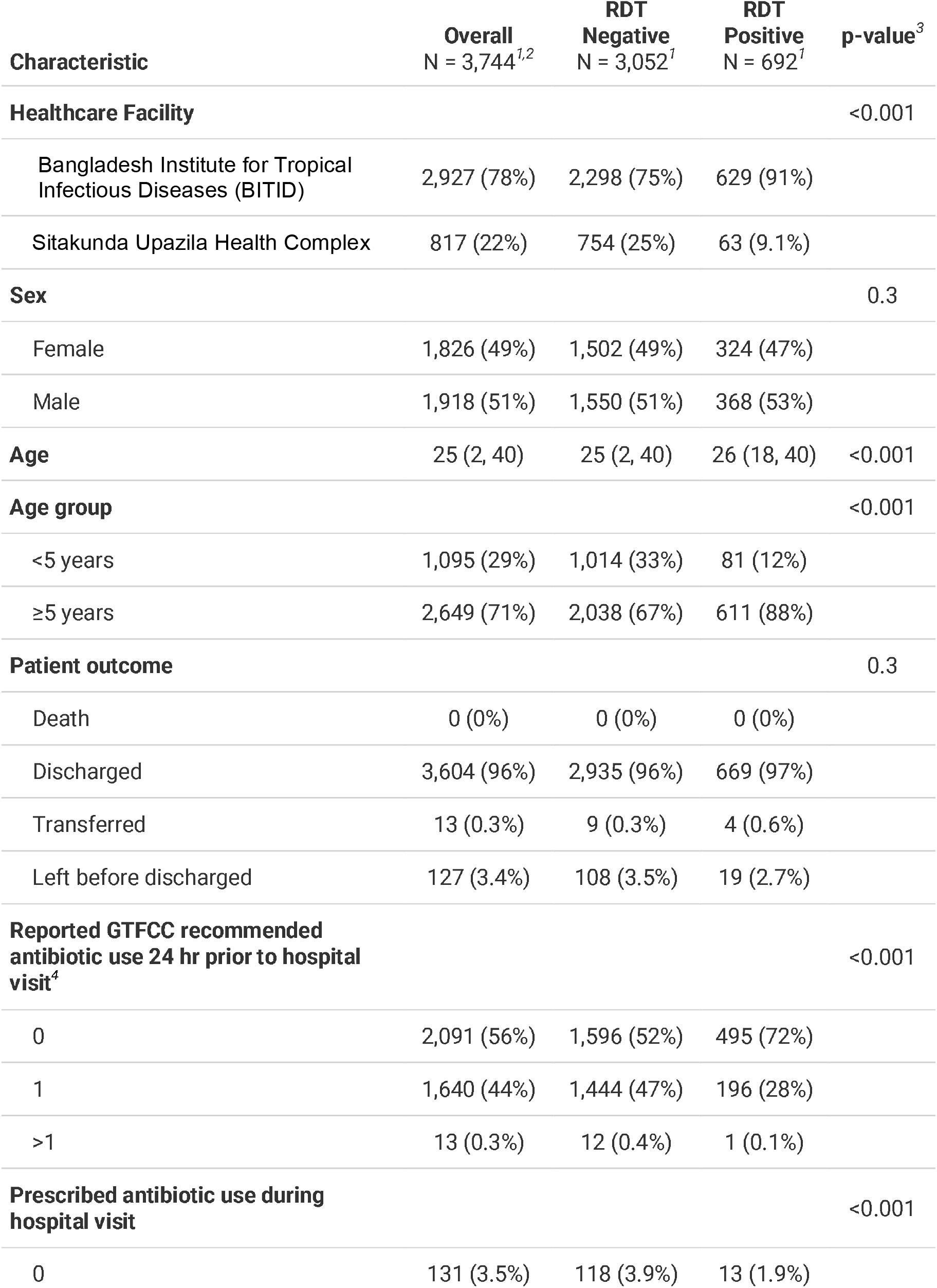
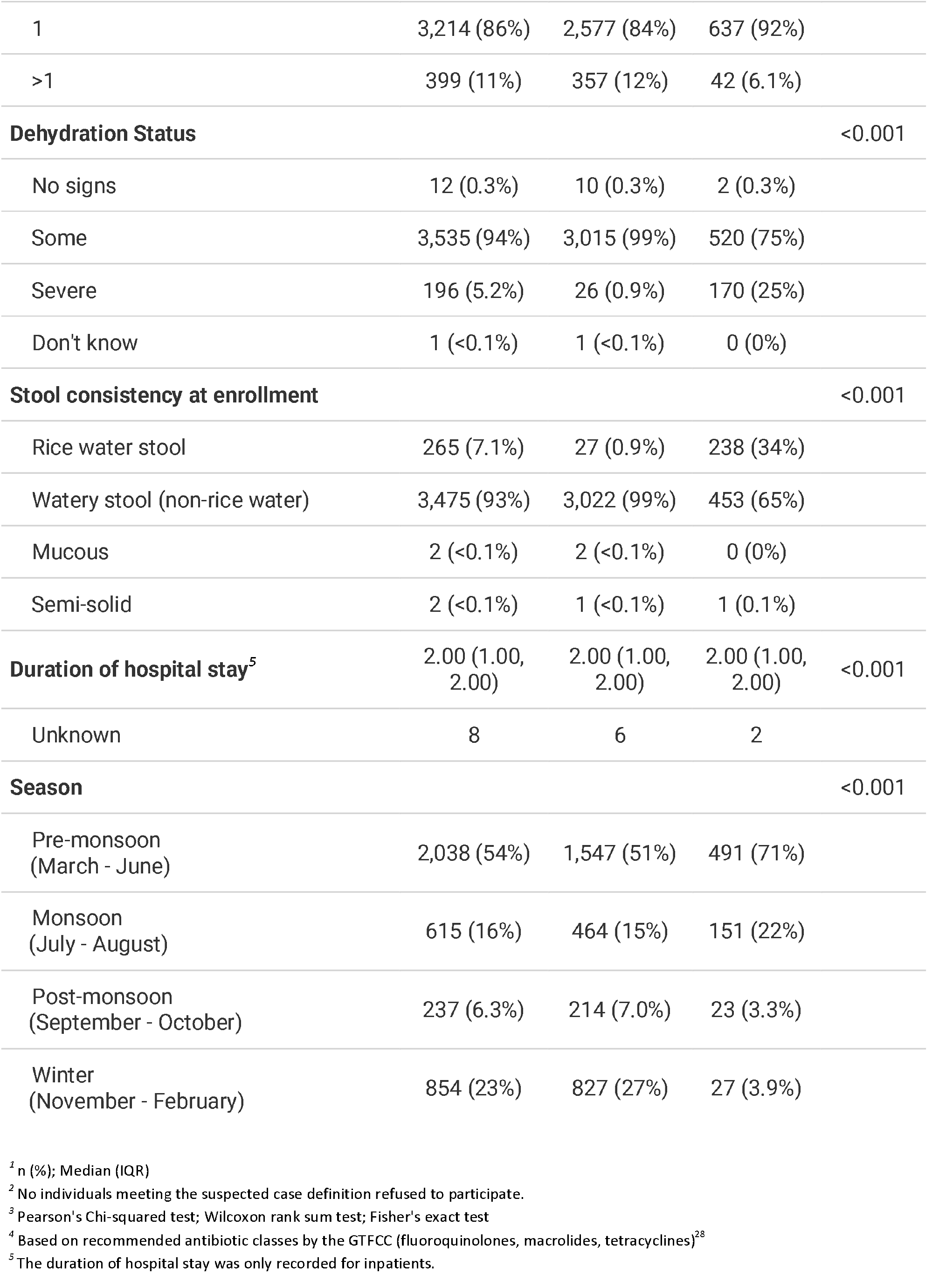
Description of participants from the start of the study until August 31, 2022.

**Figure 1:**
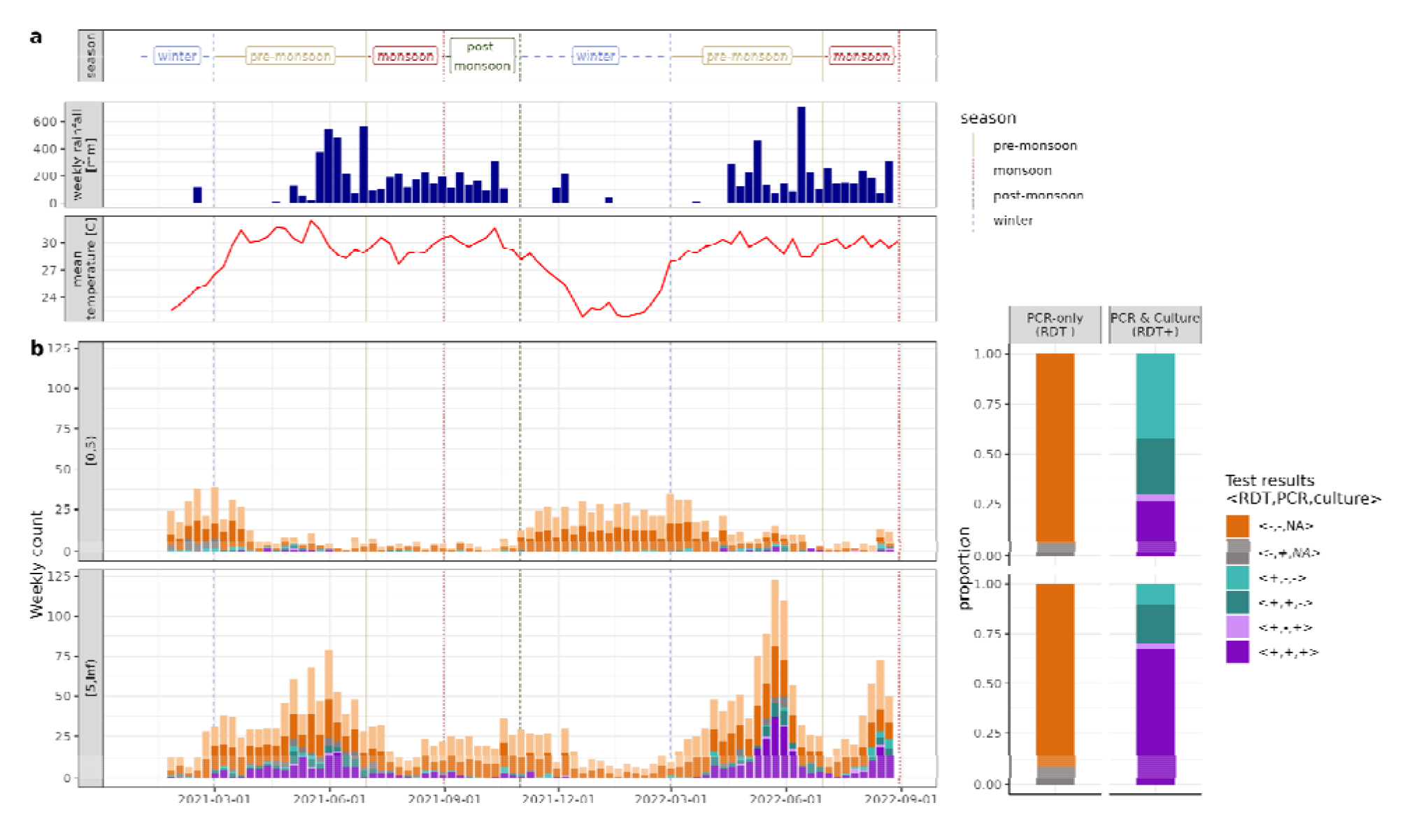
Cholera surveillance diagnostic results. a) Definition of seasons (winter, pre-monsoon, monsoon, post-monsoon) and weather in terms of weekly precipitation (Imerg dataset https://gpm.nasa.gov/data/imerg) and weekly mean temperature (recorded at Shah Amanat International Airport in Chattogram downloaded from https://bmd.gov.bd/link/aws). b) Right: Weekly counts by 3-way category of RDT (Cholkit), PCR and culture test results, stratified by age category. Left: Proportion by 3-way test category for samples with PCR or PCR and culture results, stratified by availability of culture results. Due to our sampling protocol, only around 50% of RDT-samp es were tested by PCR (left column, N=1391), and all RDT+ samples were systematically tested by PCR and culture (right column, N=691).

The number of RDT-positive tests and PCR confirmation rates also varied strongly by season, with few RDT-positive tests being reported during the winter (November-February) and a low proportion of those being PCR-confirmed (37.0%, 10/27), compared to most RDT-positive tests being reported during the pre-monsoon period (March-June) with high PCR confirmation rates (88.8%, 435/490). In univariate analysis, positive RDT tests were more likely to occur among participants older than 5 years, during the pre-monsoon and monsoon periods, presenting with severe dehydration, without prior use of antibiotics and with rice watery stool (Table 1).

After accounting for time-varying cholera incidence rates and partial testing, we estimate a sensitivity of 93.4% (95% CrIs: 91.3-95.3) for RDT, 90.3% (88.4-92.1) for PCR, and 73.7% (70.8-76.6) for culture, and a specificity of 97.3% (96.7-97.8) for RDT, and 97.2% (96.6-97.8) for PCR (Table 2, culture specificity was assumed to be 100%) (Figure 2). However, these mean estimates mask significant variations across patient and sampling characteristics. In regression models that account for possible confounding (Supplementary Figure S1), we found statistically significant effects of age group on the sensitivity of all three tests, in addition to antibiotic use and time to sample processing for culture (odds ratios for all covariate in Supplementary Figure S3). The sensitivity of culture and PCR was lower for children under 5, especially for culture where the post-stratified mean was 42% (95% CrI: 31-54) compared to 72% (95% CrI: 69-76) for those older than 5 years. Culture sensitivity was also lower among those reporting to have taken a GTFCC-recommended antibiotic (56%; 95% CrI: 49-63) compared to those who did not (69%; 95% CrI: 65-73), and by the delay from sample collection to testing. Sensitivity of culture reduces from 82% (95% CrI: 78-87) when performed on the day of collection, to 65% (95% CrI: 60-70) after 2 weeks to 54% (43-65) after 3 weeks. Finally, the strongest seasonal differences were between PCR sensitivity in the pre-monsoon and the monsoon periods (96% [95% CrI: 94-97] vs. 82% [95% CrI: 78-86]). We found less pronounced variations in test specificity, with only season having a statistically significant effect on RDT and PCR, but estimates were consistently found to be above 95%, with the lowest values during the monsoon period.

**Table 2.** CholKit RDT performance. Unadjusted estimates are computed using PCR as gold standard, as culture was only performed on RDT+ samples. Model-adjusted estimates account for imperfect test performance as well as changes in underlying cholera incidence.

| Metric | N reference PCR results | N discordant RDT results | Unadjusted estimate (95% CIs) | Model-adjusted estimate (95% CIs) |
| --- | --- | --- | --- | --- |
| Sensitivity | 684 (PCR+) | 111 (RDT-) | 83.8 (80.8-86.3) | 93.3 (91.0-95.3) |
| Specificity | 1,398 (PCR-) | 118 (RDT+) | 91.6 (90.0-92.9) | 97.2 (96.7-97.7) |

**Figure 2:**
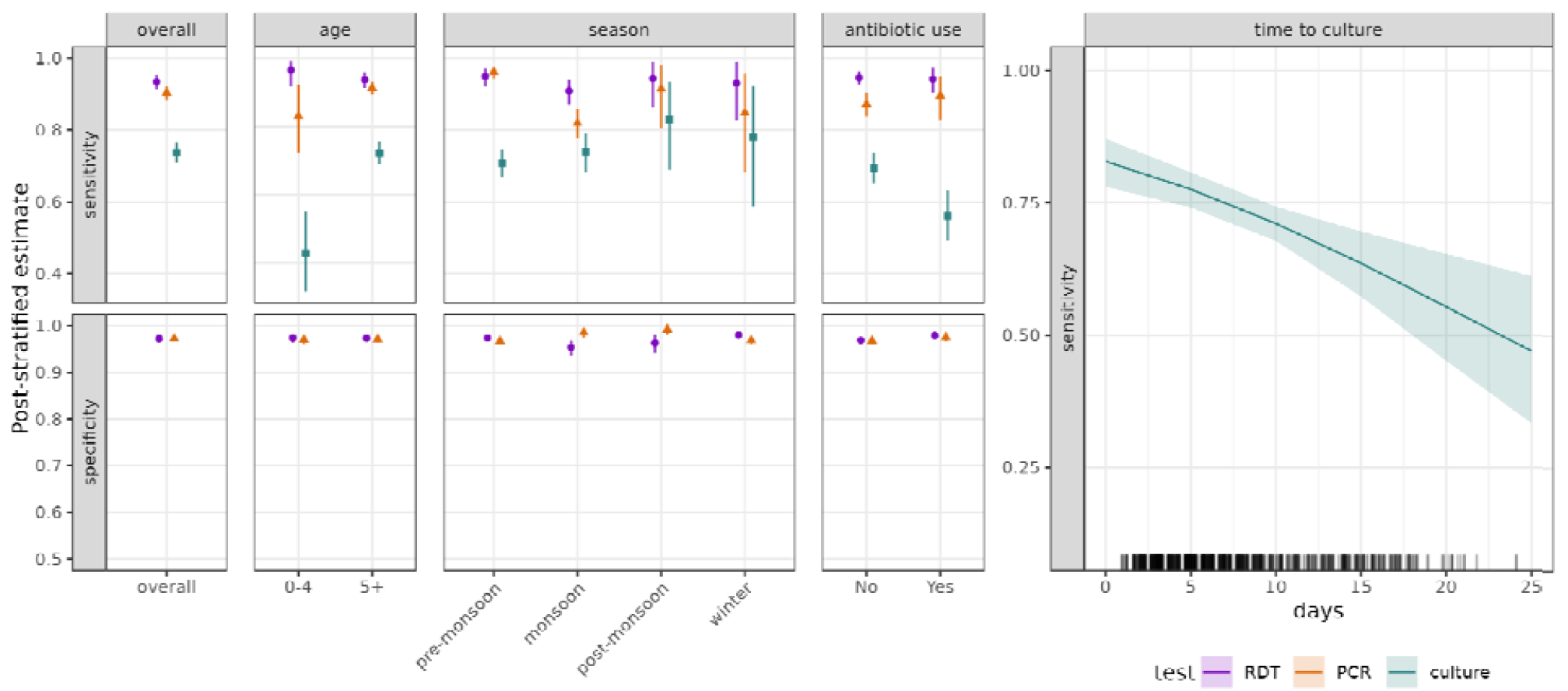
Cholera diagnostic test performance overall and by individual- and setting-specific factors. a) Overall model estimates of RDT, PCR and culture sensitivity and specificity. b) Effect of selected covariates on diagnostic test performance in terms of the odd-ratio mean (dots) and 95% CrIs (bars) of a positive test result conditional on cholera presence (top row, sensitivity), and of a negative test result condition on cholera absence (bottom row, specificity). Note that culture specificity is assumed perfect and therefore not modeled. Post-stratified estimates (dots: mean, bars: 95% CrIs) for each discrete covariate strata (age, season, antibiotics), and continuous covariate values (time to culture).

Although we find that participant-level and sampling factors may affect test performance, changes in the underlying prevalence of cholera among tested AWD cases can also have important implications for RDT result interpretation. Accounting for age-specific changes in seasonal cholera prevalence among AWD cases as well as variations in RDT performance illustrates that the negative predictive value (NPV) of RDT was consistently high in our study (above 98%), but that the positive predictive value (PPV) varied strongly (Figure 3). For children under 5, the PPV was lowest during the winter at only 14% (95% CrI: 11-18) when the mean cholera prevalence among AWD cases was lowest (0.5%), and highest during the monsoon period at 82% (78-86) when cholera prevalence was 11%. The PPV was consistently higher in the older age class, ranging from 55% (45-66) in the winter with 3% cholera prevalence, to 93% (90-96) in the pre-monsoon period when prevalence was 27%. Due to lower sensitivity, the NPV of culture was systematically lower than that of RDT, including an average NPV of less than 85% during the pre-monsoon season among those aged five years and older (Supplementary Figure S6).

**Figure 3:**
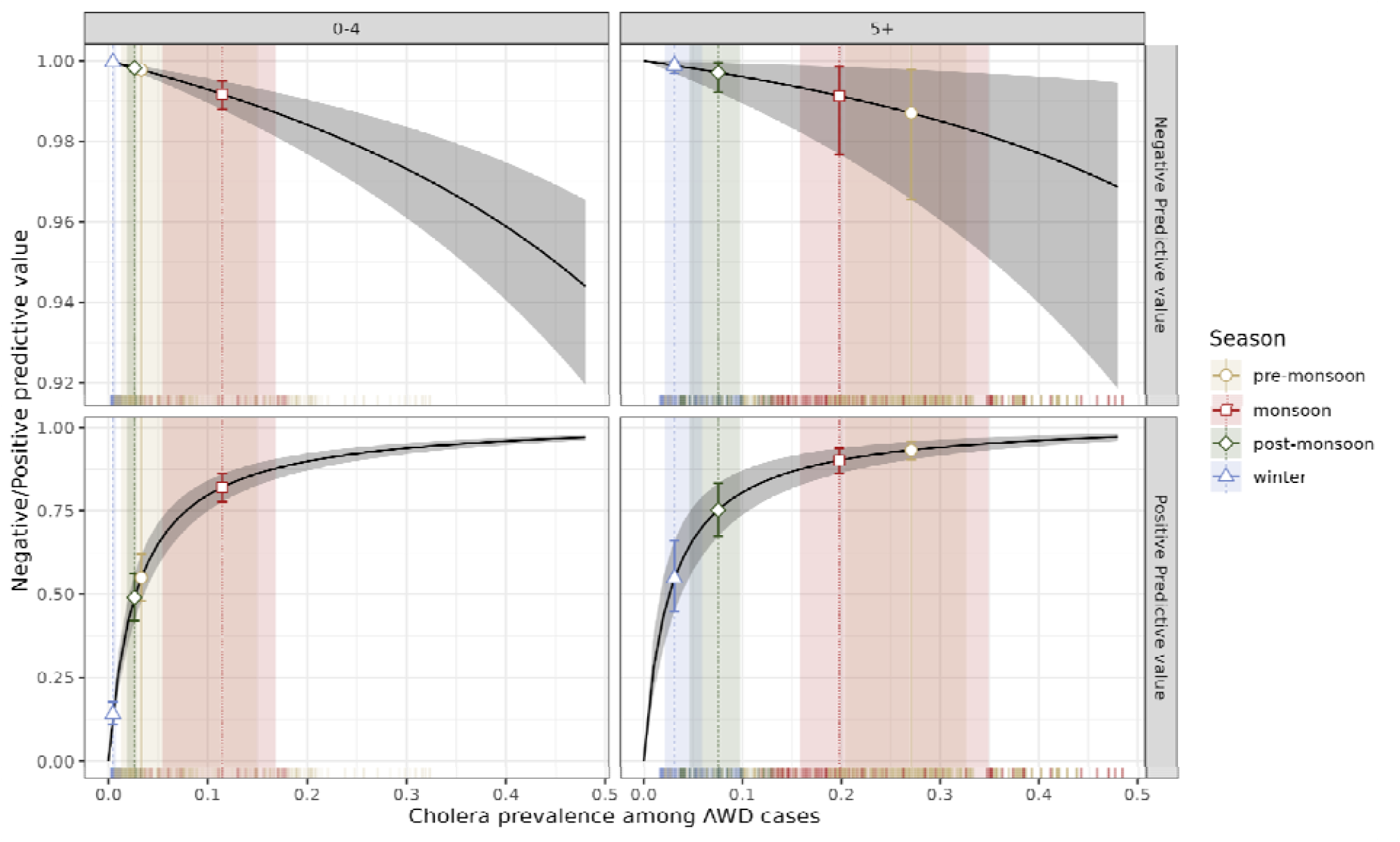
Rapid diagnostic test negative and positive predictive values in study. Negative (NPV) and positive (PPV) predictive values for Cholkit RDT across age classes (columns) and seasons in Sitakunda, Bangladesh (colors). Black lines indicate NPV/PPV values computed at the overall mean estimates of RDT sensitivity and specificity (Table 2), and gray ribbons indicate the 95% CrIs. Rug plots indicate the estimated prevalence of cholera among AWD cases at days with at least one AWD case by age category during the study period. Vertical lines indicate the median cholera prevalence among AWD in each season and by age category, and dots indicate the corresponding mean NPV/PPV (error bars give the corresponding 95% CrIs).

Through simulated field evaluations, we found that estimates of RDT performance where PCR, culture or a combination of both are used as reference standards may be severely under-estimated, and that the magnitude of the bias depends on the underlying cholera prevalence among AWD cases (Figure 4). Underestimation of sensitivity is greatest when cholera prevalence is low and PCR or the combination of PCR and culture are used as reference standards. For instance, under monsoon-season conditions (median cholera prevalence of 18%, interquartile range, IQR: 15-32), RDT sensitivity estimates would be 85% when compared to PCR or PCR/culture, which is lower than the ‘true’ value of 93.4% used in the simulations. In contrast, the negative bias in estimates of RDT specificity increases with increasing cholera prevalence among AWD, especially when using culture as the reference standard. For example, under monsoon-season conditions the estimates of RDT specificity would be 92% when compared to culture, which is lower than the ‘true’ value of 97%; using the composite of PCR and culture as the reference assay would have yielded the best estimate just below the ‘true’ value of 97%. Given a cholera prevalence among AWD cases of 75%, as is often observed in outbreak situations,^29^ RDT specificity would be greatly underestimated when using culture (mean RDT specificity estimate of 55%) or PCR (71%) as reference assays, with the combination of PCR and culture yielding more accurate, though still under-estimated, results (91%).

**Figure 4:**
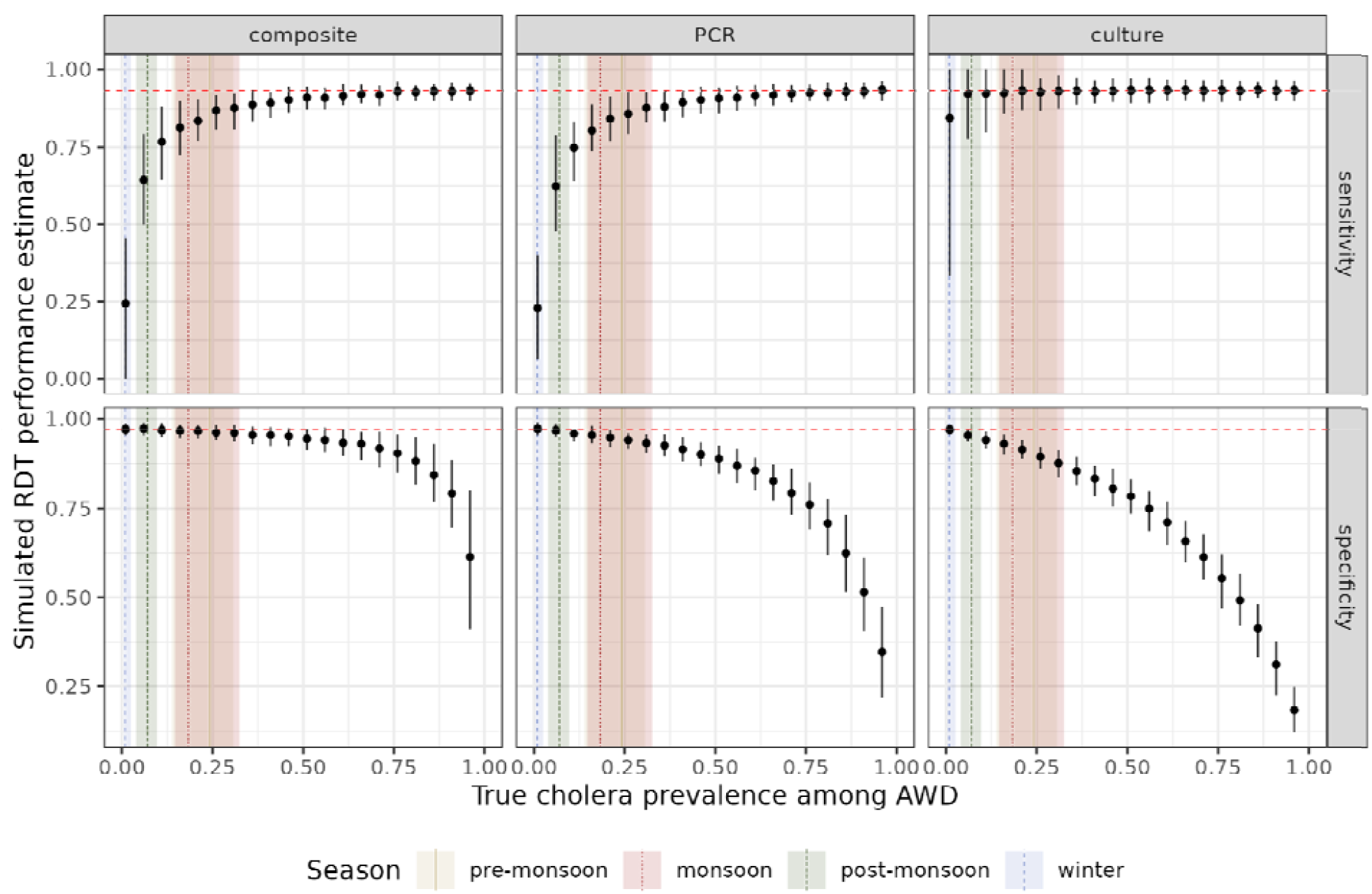
Simulations of bias in RDT evaluation due to the imperfect nature of PCR and culture. RDT performance was evaluated using 100 simulations of 300 synthetic samples when considering three different reference assay definitions (columns: composite as combination of PCR and culture, PCR and culture), across a range of true cholera frequency (5% to 95%). Horizontal dashed lines indicate the true RDT specificity and specificity values used for simulations. Alignment along the horizontal line indicates that the estimated performance aligns with that used for simulations. Dots indicate the mean and bars the 95% binomial CIs.

## Discussion

Through a direct comparison of rapid and traditional cholera diagnostics in an endemic field setting, we shed new light on test performance and the factors that can affect it. We find that the Cholkit RDT has high sensitivity (> 90%) and specificity (>95%) and illustrate how various patient- and setting-specific factors, like age, season, antibiotic use, time to testing and underlying disease prevalence, can shape not only test performance, but test result interpretation at the individual-level. Through this, we show that diagnostic field evaluations can lead to under-estimation of the sensitivity and specificity of new tests due to the imperfect nature of reference assays and that the magnitude of bias depends on the epidemiologic setting.

Though the use and availability of cholera RDTs have increased in the last decade,^6,30^ concerns of product quality and variability in field performance have hindered their wide-scale deployment. Our results suggest good performance of RDTs like Cholkit in an endemic setting, similar to PCR and consistent with previously published estimates for Cholkit RDT.^25^ With comparatively little variability across the different patient- and setting-specific factors observed in our study, these results support the use of RDTs within cholera surveillance programs and for probable outbreak detection, particularly when confirmatory testing is inaccessible.

Culture has historically served as the reference diagnostic for confirming cholera, declaring outbreaks and evaluating novel diagnostics. Our findings add to the existing evidence that culture sensitivity is moderate to low (<75%),^25^ with drastically reduced sensitivity among children under five (<45%), which has not previously been documented.^12,31^ As our age-specific estimates control for antibiotic use, lower bacterial concentrations or different co-circulating pathogens that interfere with the culturability of *V. cholerae* in-vivo may render culture less sensitive among children in this setting. This differential sensitivity of culture could have profound consequences on our interpretation of results from many seminal studies, including randomized trials of oral cholera vaccines, that were based on culture.^32^ Our analysis further confirms that, in addition to age, antibiotic use prior to visiting the health facility greatly reduces culture sensitivity (∼ 13%).^16–18,21,33^ Importantly, we find that delayed testing may strongly increase false negative rates, with negative predictive values going from 95% to 87% in a typical pre-monsoon season when testing samples three weeks after collection. These results suggest that the timely analysis of samples, age, and antibiotic use are all key for using and interpreting culture as a confirmatory assay and particular care should be given to ruling out cholera with negative tests in many situations, in contrast to general recommendations by the GTFCC.^6^

While our study was not powered to detect differences in test performance between seasons, our adjusted point estimates suggest the accuracy of cholera diagnostics may vary throughout the year. We observed reduced sensitivity of culture during pre-monsoon (high prevalence) season compared to PCR and RDT; and relatively high culture, PCR, and RDT sensitivity during post-monsoon season. While age and antibiotic use could change seasonally, these were controlled for in our model and other factors could explain differences in performance by season. Notably, the prevalence of other enteric pathogens (and non-pathogens) varies seasonally and co-infection with other organisms could inhibit concentrations and growth of *V. cholerae* O1.^34^ Lytic bacteriophages, which have a clear seasonal pattern in Bangladesh, may decrease the sensitivity of diagnostics.^15,35^ Lastly, exposure pathways to *Vibrio cholerae* O1 may vary seasonally due to changes in the availability and quality of water supplies,^36^ which could in turn impact the infectious dose, disease severity, and resulting concentration of bacteria in the stool. As previously shown, the ratio of symptomatic to asymptomatic infections changes across seasons, which could be explained by time-dependent exposure routes and infectious dose.^23^

At the individual-level, our results stress the need to account for the underlying cholera prevalence when interpreting test outcomes. The probability of true disease varies largely in time and between age classes,^29^ and as a result, the probability that a positive test result is a true positive (PPV) ranged enormously from <15% during the winter season (<1% cholera prevalence among those with AWD) among children less than 5 years old to >90% during the pre-monsoon season (∼25% cholera prevalence) among those greater than 5 years old. The context specific PPV of cholera RDTs could, in some situations, allow us to have high confidence in positive test results for both outbreak alerts and individual-level diagnosis, a consideration for future global surveillance recommendations.^6^

The context in which a RDT (or other new diagnostic test) is evaluated matters, and the underlying disease prevalence is a key variable when evaluating RDT performance due to the imperfect nature of PCR and culture. Simulations show that RDT specificity will tend to be underestimated when evaluated against culture and PCR in outbreak settings when cholera prevalence among AWD cases is high (> 50%) with sensitivity being underestimated when cholera prevalence is low (< 25%). Use of a composite PCR/culture reference assay may mitigate these biases, but do not correct for them completely. These results, therefore, stress the importance of accounting for the setting and study population in which RDTs are evaluated, and the value of latent class models to draw inference in the absence of perfect reference assays. These considerations are not limited to the evaluation of cholera RDTs, and similar qualitative patterns can be expected in any test evaluation against an imperfect reference assay.

The strength of this study is its large sample size and our joint inference of test performance accounting for imperfect tests, partial testing and the underlying time-varying cholera incidence. However, this study also comes with some limitations. Firstly, we lacked complete diagnostic test results for all samples, particularly for RDT-negative samples, where complete culture results were not available. Nevertheless, our modeling framework allowed us to address this partial testing. Secondly, our study was conducted in an endemic setting and, thus, our overall RDT performance results and those stratified by age and season may not be generalizable to other settings.

In its entirety, this work supports the use of RDTs in cholera surveillance and an improved methodological framework for evaluating the field performance of cholera diagnostics. RDTs can offer diagnostic support at the individual-level,^37^ if interpreted correctly, and can play a critical role in surveillance for cholera control and elimination programs. Both the accuracy and interpretation of a test are important for the rational use of diagnostics. Given the imperfect performance of available reference assays, field evaluations should use multiple tests and latent class models to reduce potential biases and simultaneously assess the variability of estimates across a suite of patient- and setting-specific factors. Further, with the prevalent abuse of antibiotics, we need to reconsider how negative culture results are interpreted. Together, these results emphasize how dependent our assessment of cholera diagnostics is on context and urge us to tailor recommendations appropriately in order to make progress on the pathway towards cholera elimination.

## Data Availability

All data produced in the present study are available upon reasonable request to the authors.

https://github.com/HopkinsIDD/rethinking_cholera_diagnostics/

