## Supplemental Material for "Rethinking cholera diagnostic test performance, interpretation and evaluation: a field-based latent-class analysis in Bangladesh"

#### Contents

**S1 Modeling framework** **S2**

S1.1 Latent class model . . . . . S2

S1.2 Participant and sampling effects on test performance . . . . . S3

S1.3 Correcting for possible confounding . . . . . S3

**S2 Supplementary Figures** **S6**

S2.1 Figure S2: Venn diagrams of RDT, PCR and culture results by age . . . . . S6

S2.2 Figure S3: Inferred effect sizes . . . . . S7

S2.3 Figure S4: Post-stratified estimates by age category . . . . . S7

S2.4 Figure S5: Sensitivity analysis on antibiotics definition . . . . . S8

S2.5 Figure S6: NPV and PPV for culture only . . . . . S9

S2.6 Figure S7: Posterior retrodictive checks . . . . . S10

**S3 Supplementary Tables** **S11**

S3.1 Age distribution of study participants . . . . . S11

S3.2 Antibiotic use among study participants . . . . . S11

S3.3 Antibiotic class among participants reporting antibiotic use . . . . . S12

### S1 Modeling framework

The aim of the modeling framework was to estimate the infer the performance of Cholkit RDT, PCR and culture as well as the impact of patient-level characteristics and sampling factors. To do so we develop a latent class model that accounts for imperfect tests, changes in underlying cholera prevalence among AWD cases, and the specificity of our sampling protocol. We also account for possible confounding between covariates to estimate causal effects.

#### S1.1 Latent class model

##### S1.1.1 Probability of true cholera

For each enrolled patient presenting with AWD we model the prior probability $\phi_i(t)$ of having cholera. This probability accounts both for the estimate of the time-varying changes in the age-class specific population-level cholera prevalence among AWD case, $\rho_a(t)$, where $a$ is the age class of patient $i$, and participant-level characteristics:

$$\log(\phi_i(t)) = log(\rho_a(t)) + \boldsymbol{\beta}_{chol}\boldsymbol{X}_i + \gamma T(t),$$

where matrix $\boldsymbol{X}_i$ encodes participant characteristics, here age and dehydration status at admission, and $T(t)$ is the mean weekly temperature as measured in the Shah Amanata International Airport in Chattogram.

We then connect the results of RDT, PCR and culture to the patient-level probability of having cholera through the likelihood of observing test results given the underling state.

##### S1.1.2 Accounting for multiple tests

The surveillance scheme implemented in the study is described in details in Hegde et al. (2024) Briefly, all suspected cholera cases were tested with RDT¸ around half of RDT-negatives were tested with PCR, and all RDT-positives were tested with PCR and culture.

If all test test had been performed, the likelihood of the data would correspond to a multinomial distribution:

$$\begin{aligned}&\left[n_{\{-,-,-\}}, n_{\{-,-,+\}}, n_{\{-,+,-\}}, n_{\{-,+,+\}}, n_{\{+,-,-\}}, n_{\{+,-,+\}}, n_{\{+,+,-\}}, n_{\{+,+,+\}}\right] \sim \\ &multinomial(p_{\{-,-,-\}}, p_{\{-,-,+\}}, p_{\{-,+,-\}}, p_{\{-,+,+\}}, p_{\{+,-,-\}}, p_{\{+,-,+\}}, p_{\{+,+,-\}}, p_{\{+,+,+\}}),\end{aligned}$$

where signs in brackets indicate the result of RDT, PCR and culture respectively (e.g., $\{-,+,-\}$ indicates a negative RDT, a positive PCR and a negative culture result). The vector $p$ is the probability of a test outcome accounting both for the probability of cholera $\phi$ (which is be participant and time-specific as detailed above), and test sensitivity $\theta^+$ and specificity $\theta^-$:

$$\begin{aligned}
p_{\{-,-,-\}} =&(1-\theta_1^+)(1-\theta_2^+)(1-\theta_3^+)\phi + \theta_1^-\theta_2^-\theta_3^-(1-\phi),\\
p_{\{-,-,+\}} =&(1-\theta_1^+)(1-\theta_2^+)\theta_3^+\phi + \theta_1^-\theta_2^-(1-\theta_3^-)(1-\phi),\\
p_{\{-,+,-\}} =&(1-\theta_1^+)\theta_2^+(1-\theta_3^+)\phi + \theta_1^-(1-\theta_2^-)\theta_3^-(1-\phi),\\
p_{\{-,+,+\}} =&(1-\theta_1^+)\theta_2^+\theta_3^+\phi + \theta^-(1-\theta_2^-)(1-\theta_3^-)(1-\phi),\\
p_{\{+,-,-\}} =&\theta_1^+(1-\theta_2^+)(1-\theta_3^+)\phi + (1-\theta_1^-)\theta_2^-\theta_3^-(1-\phi),\\
p_{\{+,-,+\}} =&\theta_1^+(1-\theta_2^+)\theta_3^+\phi + (1-\theta_1^-)\theta_2^-(1-\theta_3^-)(1-\phi),\\
p_{\{+,+,-\}} =&\theta_1^+\theta_2^+(1-\theta_3^+)\phi + (1-\theta_1^-)(1-\theta_2^-)\theta_3^-(1-\phi),\\
p_{\{+,+,+\}} =&\theta_1^+\theta_2^+\theta_3^+\phi + (1-\theta_1^-)(1-\theta_2^-)(1-\theta_3^-)(1-\phi),
\end{aligned}$$

where subscripts 1,2,3 denote RDT, PCR and culture, respectively.

Due to our sampling protocol, we do not have test results of PCR and culture for certain participants. To account for partial testing, we follow our previous approach which factors in the conditional probabilities of unobserved PCR and culture conditional on the result of RDT. We refer the reader to our Hegde et al. (2024) for further details (Methods, section "Statistical analysis").

#### S1.2 Participant and sampling effects on test performance

We here account for the impact of participant characteristics (age, antibiotic use) and sampling (RDT batch, season, time to culture) on test performance by incorporating them as covariates in our latent class model. Specifically, we assume that the sensitivity, $\theta^+$, and specificity, $\theta^-$, of RDT, PCR, and culture follow a logit-linear model:

$$logit(\theta_j^+) = \beta_{j,0}^+ + \boldsymbol{\beta}_j^+ \boldsymbol{\mathcal{X}}_j^+,$$
$$logit(\theta_j^-) = \beta_{j,0}^- + \boldsymbol{\beta}_j^- \boldsymbol{\mathcal{X}}_j^-,$$

where $j$ is the test index as above (1: RDT, 2:PCR, 3:culture), $\beta_{j,0}^{+/-}$ is the intercept, $\boldsymbol{\beta}_j^{+/-}$ is the vector of covariate coefficients, and $\boldsymbol{\mathcal{X}}_j^{-/+}$ is the covariate matrix.

#### S1.3 Correcting for possible confounding

The relationship between participant characteristics, sampling factors and test performance may be complex and presents the challenge of estimating effects in the presence of possible confounding between factors. To address this we propose a directed acyclical graph representing our assumptions on how these measurable factors are causally connected through unmeasurable quantities which ultimately may impact the performance of RDT, PCR and culture (Figure S1).

Given each DAG and covariate, we define the set of covariates to control for possible confounding. We do so automatically using the dagitty package in R. The final set of equations used for each factor and covariate is given in Table S1.

To account for possible RDT batch effects, following our previous analysis we separate the modeling period into two distinct periods, one from the start of the study up to June 29th 2021, and one from June 30th to the end of the study period (Hegde et al. 2024).

Table S1: Regression equations to control for possible confounding based on DAGs.

| | Test characteristic | |
|---|---|---|
| covariates | sensitivity | specificity |
| **RDT** | | |
| age | age + antibiotic use + RDT batch | age |
| antibiotics | antibiotic use + age + RDT batch | antibiotic use + age |
| batch | RDT batch + age + season | |
| period | season + RDT batch | season |
| time to culture | | |
| **PCR** | | |
| age | age + antibiotic use | age + RDT batch |
| antibiotics | antibiotic use + age | antibiotic use + age + RDT batch |
| batch | | |
| period | season | season + RDT batch |
| time to culture | | |
| **culture** | | |
| age | age + antibiotic use + time to culture | |
| antibiotics | antibiotic use + age + time to culture | |
| batch | | |
| period | season + time to culture | |
| time to culture | time to culture | |

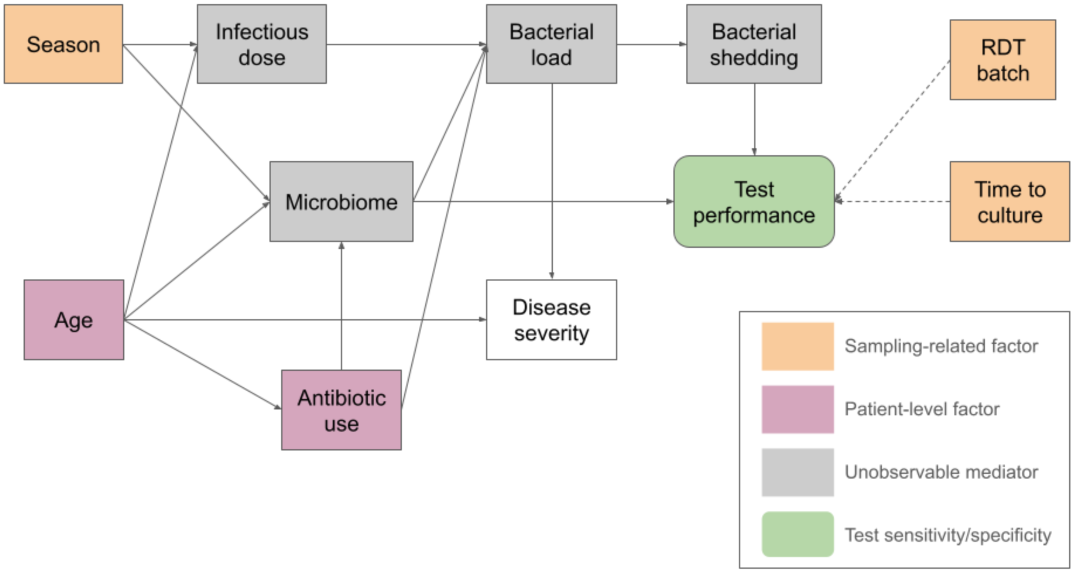


Figure S1: Directed acyclical graph of causal relations between patient-level and sampling fractors and test performance. Full arrows into test performance indicate that they affect the sensitivity of all tests. Dotted lines indicate factors that only affect certain test (RDT batch affects RDT sensitivity, and time to culture affects culture sensitivity).

##### S1.3.1 Priors

We use the following priors in the cholera incidence model with no differences between age classes:

$$
\begin{aligned}
\beta_{chol} &\sim \mathcal{N}(0, .5) \\
\gamma &\sim \mathcal{N}(0, .5) \\
\beta^{+}_{RTD,0} &\sim \mathcal{N}(0, 0.75), \\
\beta^{+}_{PCR,0} &\sim \mathcal{N}(1.04, 0.34), \\
\beta^{+}_{culture,0} &\sim \mathcal{N}(0.89, 0.26), \\
\beta^{-}_{RTD,0} &\sim \mathcal{N}(3.48, 0.69), \\
\beta^{-}_{PCR,0} &\sim \mathcal{N}(3.48, 0.44), \\
\beta^{+/-}_{i} &\sim \mathcal{N}(0, 1),
\end{aligned}
$$

,

where $\beta^{+/-}_{i,0}$ indicates the intercept for each test performance on the logit scale, $\beta^{+/-}_{i}$ the regression coefficients of the effect of covariates on test performance, and $\beta_{chol}$ the regression coefficients of the probability of true cholera. Priors for the intercepts followed our previous work in Hegde et al. (2024) based on results in Sayeed et al. (2018).

#### S2 Supplementary Figures

##### S2.1 Figure S2: Venn diagrams of RDT, PCR and culture results by age

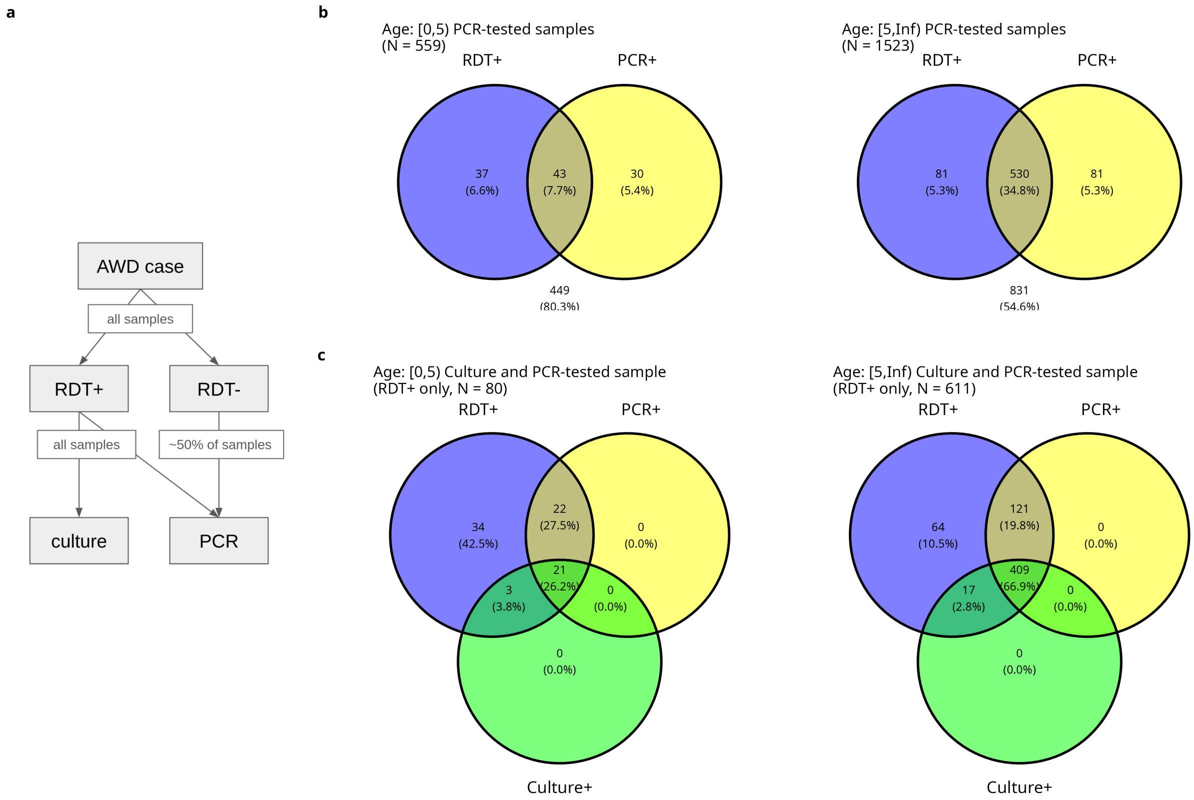


Figure S2: Raw test results. a) Study sampling scheme. All AWD cases in the study health centers were tested with RDT. All positive RDT samples were also tested with PCR and culture. Around half of RDT negative samples were tested with PCR. b) Venn diagrams of RDT and PCR test results by age class for all PCR-tested samples. c) Venn diagrams of RDT, PCR and culture test results by age class for all RDT-positive samples.

#### S2.2 Figure S3: Inferred effect sizes

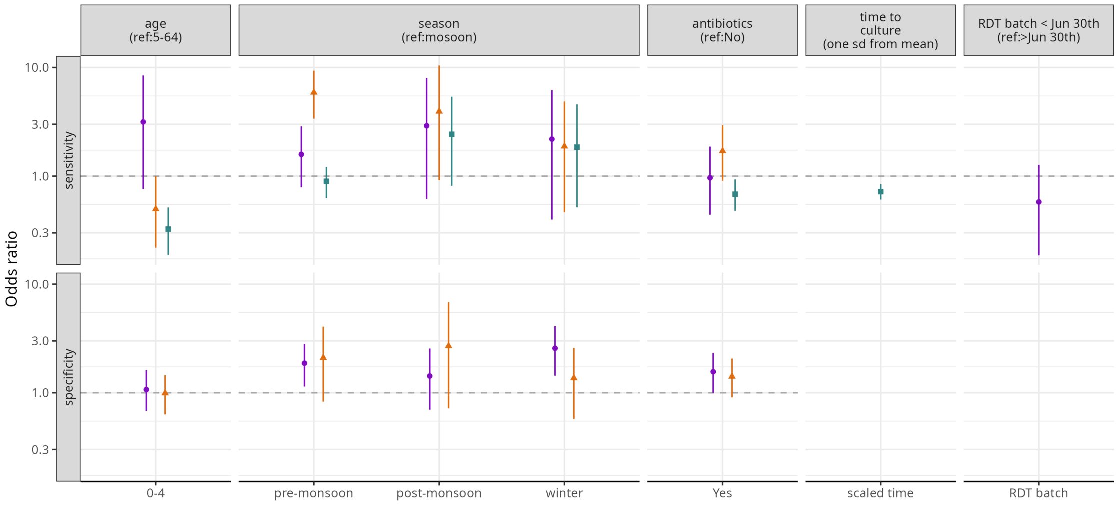


Figure S3: Covariate effect sizes on test performance. Inference of effect sizes accounts for possible confounding as described in section S2.3. Dots indicate mean of 5000 posterior HMC draws, and bars the 95% CrIs.

#### S2.3 Figure S4: Post-stratified estimates by age category

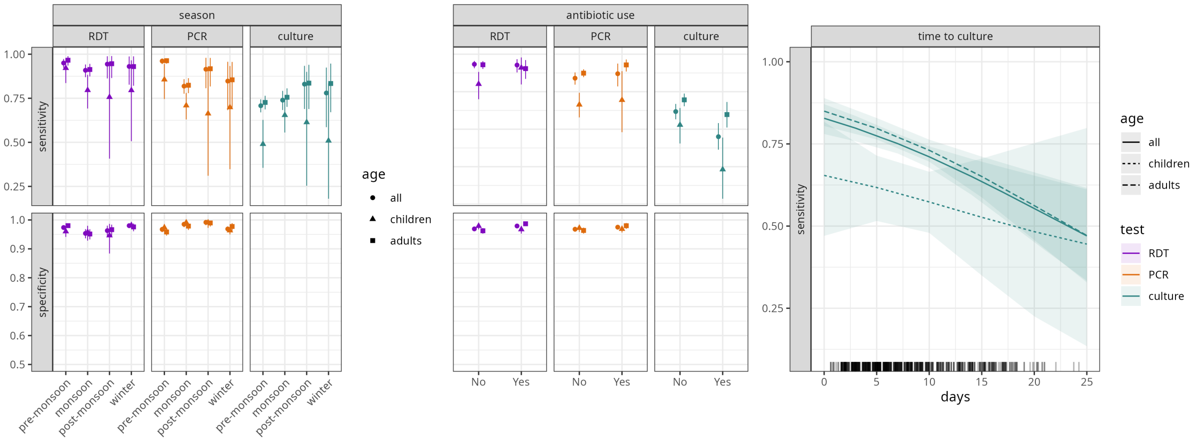


Figure S4: Post-stratified estimates by age class. Legend as in main Figure 2. Dots indicate the mean of 5000 posterior HMC draws, and bars the 95% CrIs.

#### S2.4 Figure S5: Sensitivity analysis on antibiotics definition

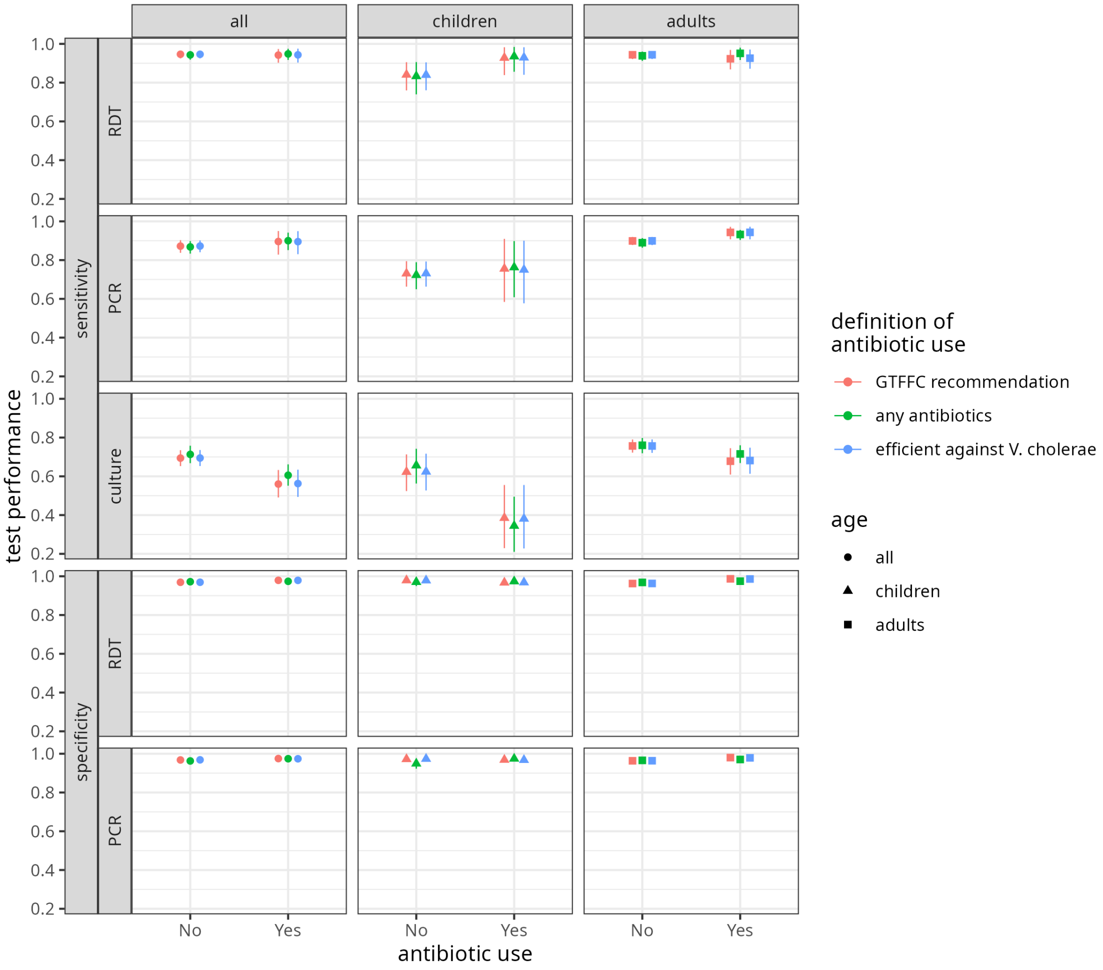


Figure S5: Post-stratified estimates for alternative definitions of antibiotic use. In addition to any antibiotic use, we grouped antibiotics either by whether they are recommended antibiotic classes by the GTFCC (tetracyclines, fluoroquinolones, and macrolides), or known to be effective against V. cholerae (fluoroquinolones, macrolides, tetracyclines, penicillins, sulfonamides, cephalosporins). We report the GTFCC-recommedended results in the main.

#### S2.5 Figure S6: NPV and PPV for culture only

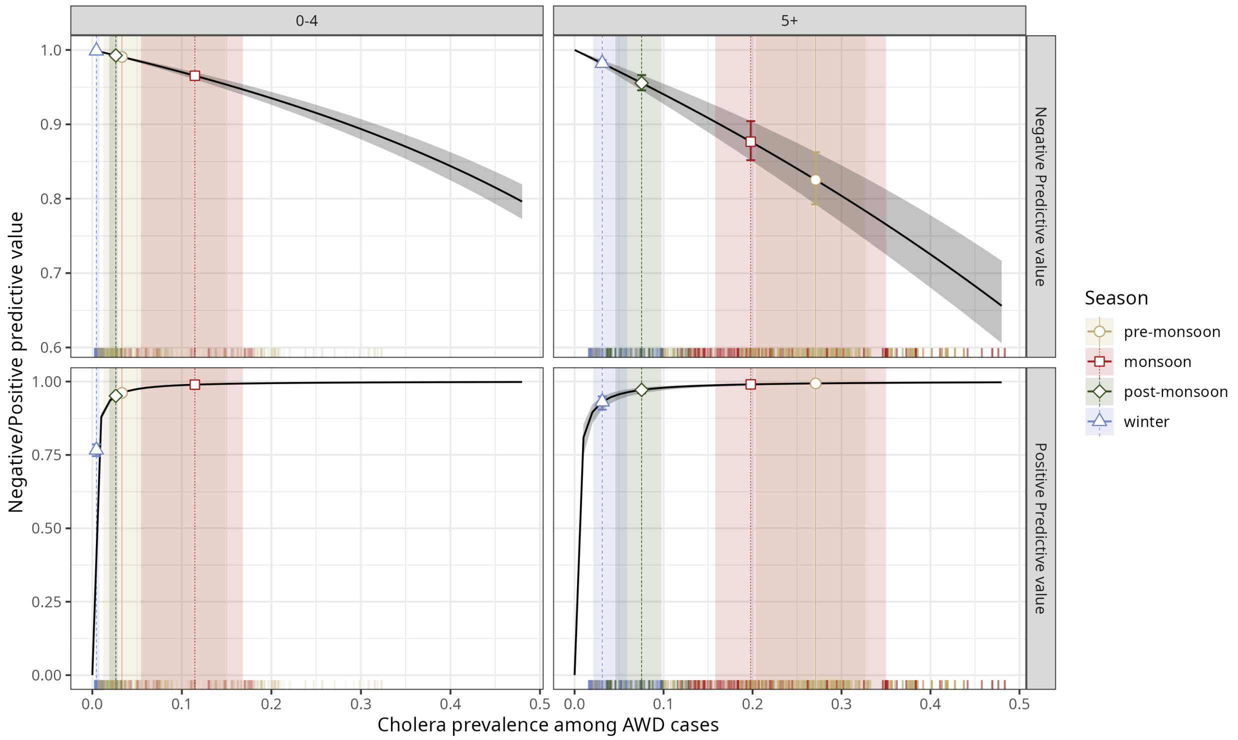


Figure S6: Negative and positive predictive values when using culture.

#### S2.6 Figure S7: Posterior retrodictive checks

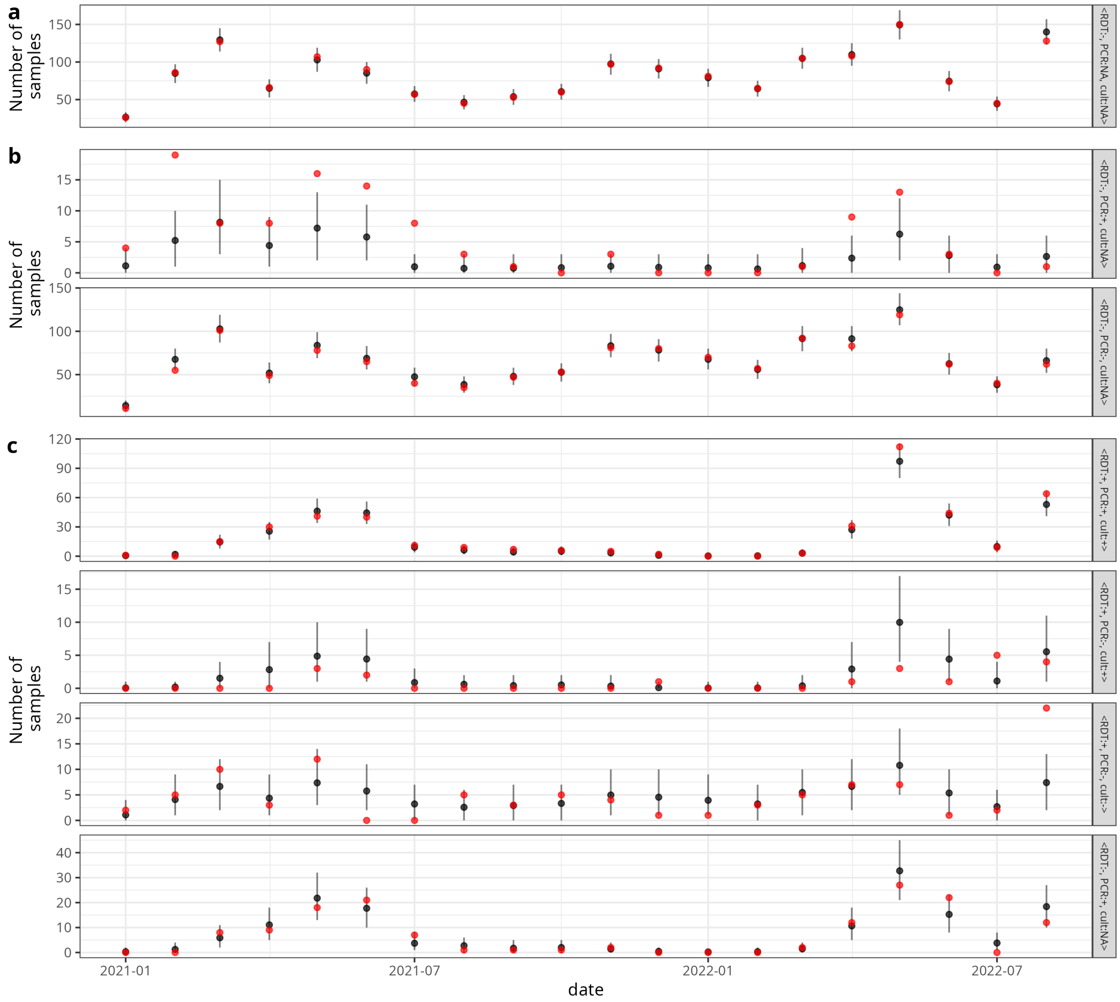


Figure S7: Posterior retrodictive checks of surveillance data. Red dots indicate monthly observed counts. Black dots indicate the mean of 5000 HMC posterior draws, and error bars the 95% CrI. a) Samples with RDT negative results only. b) Samples with both RDT and PCR result, but no culture. c) RDT positive samples with PCR and culture results.

### S3 Supplementary Tables

#### S3.1 Age distribution of study participants

Table S2: The age distribution of the study population in Sitakunda, Bangladesh.

| Age category | Overall, N = 3,744 | RDT-negative, N = 3,052 | RDT-positive, N = 692 |
|---|---|---|---|
| [0,5) | 1,095 (29%) | 1,014 (33%) | 81 (12%) |
| [5,10) | 125 (3.3%) | 87 (2.9%) | 38 (5.5%) |
| [10,15) | 65 (1.7%) | 47 (1.5%) | 18 (2.6%) |
| [15,25) | 516 (14%) | 350 (11%) | 166 (24%) |
| [25,35) | 636 (17%) | 495 (16%) | 141 (20%) |
| [35,45) | 487 (13%) | 386 (13%) | 101 (15%) |
| [45,55) | 392 (10%) | 321 (11%) | 71 (10%) |
| [55,65) | 252 (6.7%) | 210 (6.9%) | 42 (6.1%) |
| [65,75) | 136 (3.6%) | 113 (3.7%) | 23 (3.3%) |
| [75,85) | 36 (1.0%) | 26 (0.9%) | 10 (1.4%) |
| [85,Inf) | 4 (0.1%) | 3 (¡0.1%) | 1 (0.1%) |

#### S3.2 Antibiotic use among study participants

Table S3: Antibiotic use by RDT positivity among all study participants: those that took any type of antibiotic, those that took antibiotics recommended by the GTFCC (tetracyclines, fluoroquinolones, macrolides), and those that took antibiotics that are known to be effective against killing V. cholerae (fluoroquinolones, macrolides, tetracyclines, penicillins, sulfonamides, cephalosporins).

| Type of antibiotic use | Overall, N = 3,744 | RDT-negative, N = 3,052 | RDT-positive, N = 692 |
|---|---|---|---|
| **All reported antibiotic use 24hr prior to hospital visit** | | | |
| 0 | 1,181 (32%) | 878 (29%) | 303 (44%) |
| 1 | 1,855 (50%) | 1,559 (51%) | 296 (43%) |
| 1+ | 708 (19%) | 615 (20%) | 93 (13%) |
| **Reported GTFCC recommended antibiotic use 24hr prior to hospital visit** | | | |
| 0 | 2,091 (56%) | 1,596 (52%) | 495 (72%) |
| 1 | 1,640 (44%) | 1,444 (47%) | 196 (28%) |
| 1+ | 13 (0.3%) | 12 (0.4%) | 1 (0.1%) |
| **Reported effective antibiotic use 24hr prior to hospital visit** | | | |
| 0 | 2,062 (55%) | 1,569 (51%) | 493 (71%) |
| 1 | 1,662 (44%) | 1,464 (48%) | 198 (29%) |
| 1+ | 20 (0.5%) | 19 (0.6%) | 1 (0.1%) |

##### S3.3 Antibiotic class among participants reporting antibiotic use

Table S4: The number of instances antibiotics were taken by study participants prior to the health facility visit by antibiotic class. Tetracyclines, fluoroquinolones, and macrolides are the recommended antibiotic classes by the GTFCC. The nitromidazole antibiotic primarily taken is metronidazole.

| | Age class | | |
|---|---|---|---|
| Antibiotic class | less than 5 | 5+ | Overall |
| Tetracycline* | 1 (0.10%) | 1 (0.04%) | 2 |
| Fluoroquinolone* | 538 (56%) | 876 (38%) | 1414 |
| Macrolide* | 101 (10.2%) | 149 (6%) | 250 |
| Nitroimidazole | 309 (32%) | 1272 (55%) | 1581 |
| Penicillin | 1 (0.10%) | 4 (0.2%) | 5 |
| Sulfonamide | 1 (0.10%) | 1 (0.04%) | 2 |
| Cephalosporin | 14 (1.5%) | 15 (0.68%) | 29 |
| Chloramphenicol | 0 (0.0%) | 1 (0.04%) | 1 |
| Total | 965 (100%) | 2319 (100%) | 3284 |
